# Proteomic Insights into Biology of Bipolar Disorder: Implications for Health Complexity and Mortality

**DOI:** 10.1101/2024.01.17.24301415

**Authors:** Paola Rampelotto Ziani, Marco Antônio de Bastiani, Pietra Paiva Alves, Pedro Henrique da Rosa, Tainá Schons, Giovana Mezzomo, Ellen Scotton, Flávio Kapczinski, Adriane R Rosa

## Abstract

Bipolar disorder (BD) is a debilitating condition associated with a high prevalence of medical comorbidities and premature mortality. This is the first study to explore, through high-throughput-omics combined with bioinformatics, molecular signatures, pathways, and main medical diseases related to different stages of BD. Blood samples from BD patients (n=10) classified into high (BD+) or poor functioning (BD-), based on functional and cognitive status, and healthy controls (n=5) were analyzed using mass spectrometry-based proteomic analysis. Bioinformatics was performed to detect biological processes, pathways, and diseases related to BD. Eight proteins exclusively characterized the molecular profile of patients with BD+ compared to HC, while 26 altered proteins were observed in the BD-group. These altered proteins were mainly enriched in biological processes related to lipid metabolism, complement system and coagulation cascade, and cardiovascular diseases; all these changes were more prominent in the BD-group. These findings may represent systemic alterations that occur with the progression of the illness and a possible link between BD and medical comorbidities. Such comprehensive understanding provides valuable insights for targeted interventions, addressing mental and physical health aspects in subjects with BD.

## 1. Introduction

Bipolar disorder (BD) is a highly disabling disease associated with elevated rates of premature mortality [1], [2]. Patients with BD died 9 years younger than the general population [3]. This occurs not only by suicide but also due to a high prevalence of medical comorbidities [4], [5]. A recent meta-analysis of mortality in subjects with BD showed that they have a twofold increased risk of dying prematurely due to medical diseases when compared to the general population [6]. The Task Force of the International Society for Bipolar Disorders also suggested a strong association between BD and cardiovascular disease, raising questions regarding the reasons for this link. Unhealthy lifestyle factors may contribute to the high rates of medical comorbidities in BD [5]. In addition to concerns about physical well-being, these risk factors lead to more severe manifestations of BD. For example, individuals with both BD and obesity are at a higher risk of encountering more severe mood symptoms, increased resistance to treatment, and cognitive and functional impairment.

Multifactorial models, including inflammation, oxidative stress, mitochondrial dysfunction, and neurotrophic factors, partially explain BD’s accelerated aging and premature mortality [7]–[9]. Despite these known mechanisms, our understanding of the systemic changes observed throughout the course of the BD remains incomplete. In this sense, one possible approach to accelerate the discovery of this complex relationship is to utilize high-throughput-omics. The proteomics combined with a computational analysis can provide a comprehensive understanding of the systemic alterations that occur in patients with BD, also allowing the exploration of multifaceted molecular mechanisms and potential therapeutic targets. Therefore, the present study aims to characterize the molecular signatures, pathways, and disease-related ones through an integrative analysis of proteomic and bioinformatics in blood samples of patients with BD compared to controls.

## 2. Materials and Methods

Outpatients (n=10) from the BD Program at the Hospital Clinic of Porto Alegre (HCPA, Brazil) were recruited considering the following criteria: (a) age>18 years, (b) fulfilled DSM-5 criteria according to the Structured Clinical Interview for DSM-5. The control group consisted of 5 healthy volunteers from the Hemocenter at HCPA with no family history of psychiatric or neurological disorders. Patients and controls with a clinical diagnosis of intellectual disability or Alzheimer’s disease, uncontrolled medical conditions, autoimmune diseases, alcohol and drug abuse or dependence, pregnancy, or lactation were excluded. Written informed consent was obtained from all participants before their inclusion. The study protocol was approved by the Institutional Review Board of HCPA (project number 20190640).

For the present study, patients were also classified into high (BD+) or poor (BD-) functioning based on their functional and cognitive status, which was assessed using the Functioning Assessment Short Test (FAST) and Cognitive Bipolar Rating Assessment (COBRA) [10]–[13]. Therefore, BD patients were classified into BD+ if they met FAST scores < 11 and COBRA < 10, while those with FAST scores greater than 40 and COBRA greater than 25 were classified into BD-.

The blood samples (10 ml) were collected in non-anticoagulant tubes from all participants and then centrifuged at 1500 g for 15 minutes at room temperature to separate the serum. The serum was aliquoted and stored at −80°C for further analysis. The most abundant proteins were removed with the Top14 Abundant Protein Depletion Mini Spin Columns following the manufacturer’s specifications (High Select™ Depletion Spin Columns - Thermo Scientific™).

Serum samples were depleted through various steps, including washing, drying, and disulfide bond rupture. After incubation and digestion, resulting peptides underwent desalting and were analyzed using high-resolution Orbitrap mass analyzers in a Dionex Ultimate 3000 RSLC nanoUPLC system. The mass spectrometry facility RPT02H/Carlos Chagas Institute - Fiocruz Paraná conducted the analysis, employing MaxQuant version 2.2.0.0 for Peptide Spectrum Match (PSM) analysis against the UniProt Human database. Specific parameters included trypsin-specific digestion, variable modifications like oxidation and protein N-acetylation, and fixed modifications like carbamidomethyl. Settings incorporated a peptide tolerance of 7 ppm, MS/MS tolerance of 0.1 Da, allowance for 2 missed cleavages, and a 1% False Discovery Rate (FDR) for both PSM and protein assignment. Relative quantification used MaxLFQ intensity values, providing a detailed analysis of the serum protein composition. To identify differentially expressed proteins, we considered an unadjusted p-value of less than 0.05 and a change in expression of at least 15%. The STRING (Search Tool for the Retrieval of Interacting Genes/Proteins) software was used to elucidate protein-protein interactions based on differentially expressed proteins identified. Also, we employed the Enrichr tool to explore and enhance our results through information derived from the KEGG pathways (Kyoto Encyclopedia of Genes and Genomes) and the DisGeNET database (adjusted p-value < 0.00001). The approach used in Enrichr allowed us to contextualize our findings in terms of metabolic pathways and genetic associations relevant to diseases. Demographic data and clinical characteristics were analyzed using one-way ANOVA using the SPSS 18 for Windows (provided by SPSS Inc. in Chicago, IL, USA). Furthermore, we used the R software package Weighted Gene Co-expression Network Analysis (WGCNA) to perform weighted correlation network analysis aspects. Module eigengenes (i.e., the first principal component of a given module’s standardized gene expression profile) are tested for correlation with clinical variables based (on COBRA score, number of medications, number of hospitalizations, and suicide attempts) on Pearson’s r [14].

## 3. Results

The demographic and clinical characteristics of the participants are summarized in Table 1. Gender homogeneity was maintained among the groups, as all participants were women. No significant differences in age between the groups were observed (p = 0,897). As expected from the selection criteria, BD-subjects had a higher mean FAST score (p < 0.001) and COBRA score (p < 0.001) when compared to BD+ and HC. Considering the number of medications used, BD-exhibited a higher number than HC (p = 0,004).

**Table 1:** Clinical and Demographic Data.

| **Variable: mean (s.d.)** | **Controls (n = 5)** | **BD+ (n = 5)** | **BD- (n = 5)** | **F** | **p-value** |
| --- | --- | --- | --- | --- | --- |
| Age, years | 50.80 (8,075) | 50.00 (13,360) | 53.00 (9,301) | 0,110 | 0,897 |
| Number of medications | 1.20 (1,643) | 2.80 (1,304) | 5.40 (1,673) | 9,361 | *0,004* |
| Number of hospitalizations | 0 | 2.20 (1,643) | 5.80 (5,541) | 3,850 | 0,051 |
| Number of suicide attempt | 0 | 0.20 (0,447) | 3.75 (4,193) | 3,920 | 0,052 |
| MADRS score | 3.40 (3,975) | 0.40 (0,548) | 27.80 (5,495)* | 73,17 | *<0.001\** |
| YRMS score | 0.80 (1,304) | 0.20 (0,447) | 7.20 (7,855) | 3,55 | 0,061 |
| COBRA score | 11.40 (9,370) | 2.40 (0,548) | 34.00 (9,670)* | 21,89 | *<0.001\** |
| FAST score | 4.80 (7,085) | 3.80 (1,789) | 44.70 (4,637)* | 105,26 | *<0.001\** |

ANOVA (followed by Tukey)
Italic values indicate of p-value (< 0.05)
\* Compared to BD+ and HC
s.d. = standard deviation; BD+ = bipolar disorder with high functioning; BD- = bipolar disorder with poor functioning; MADRS = Montgomery-Asberg Depression Rating Scale; YMRs = Young Mania Rating Scale; FAST = Functioning Assessment Short Test; COBRA = Cognitive Complaints in Bipolar Disorder Rating Scale

The present study was conducted to identify differentially expressed proteins between BD and HC based on Liquid Chromatography-Mass Spectrometry/Mass Spectrometry (LC-MS/MS) technology. In total, 3506440 MS/MS spectra were obtained from 30 LC-MS/MS runs, of which 128163 peptide sequences were identified using 1% FDR. The LC-MS/MS detected 234 proteins corresponding to the identified peptides for the 15 participants. Figure 1 presents a protein-protein interaction network generated using STRING that illustrates the interactions between proteins in the BD and HC groups.

**Figure 1:**
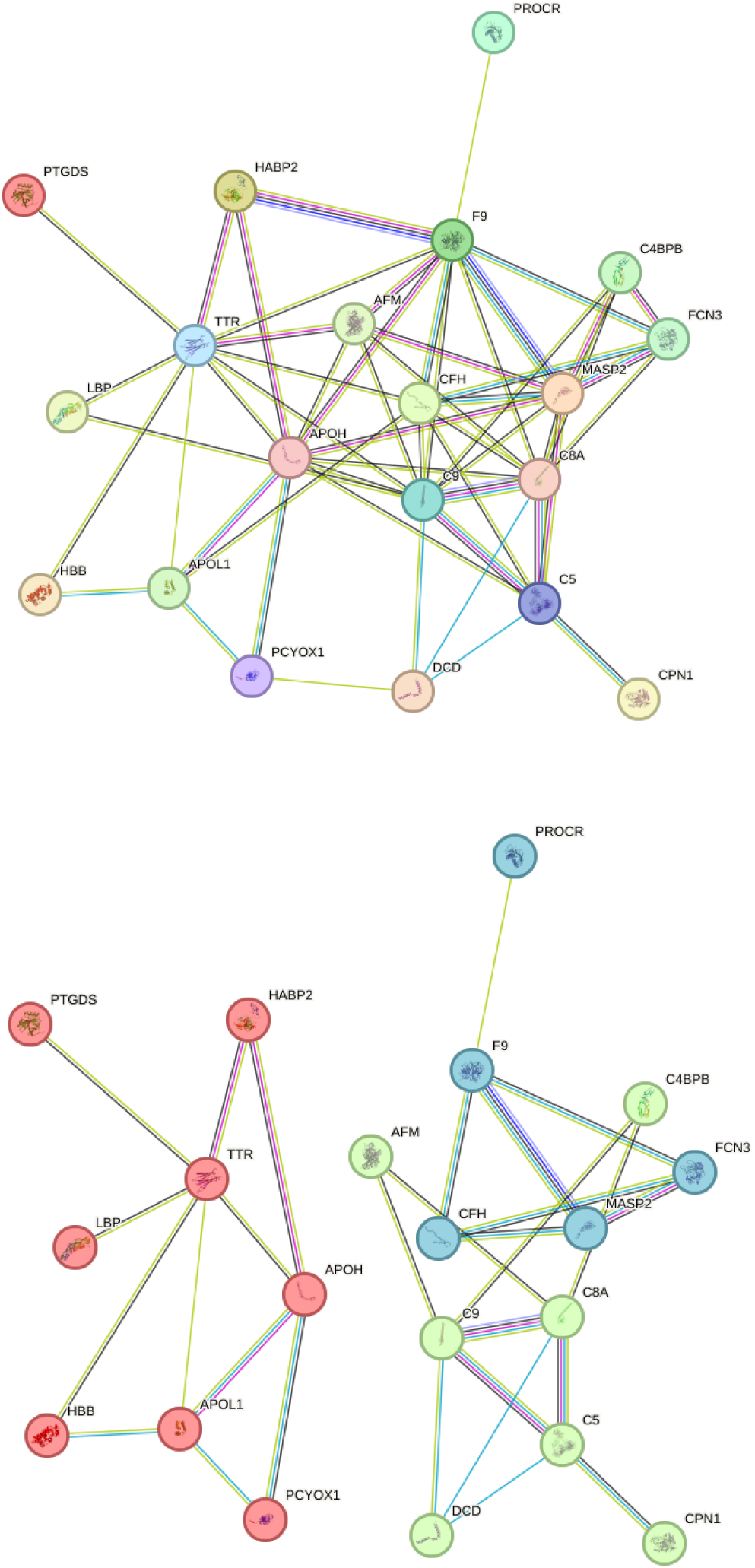
Protein-protein interaction network. The differentially expressed proteins in the BD vs HC comparison revealed the following interaction network and their respective clusters.

After statistical analysis, we identified 21 differentially expressed proteins in BD vs. HC, 12 in BD+ vs. HC, and 30 in BD-vs. HC, as shown in Table 2. The Venn Diagram displays the intersection of proteins identified in the three comparison groups. Furthermore, four proteins—C4BPB, C5, C9, and DCD—were consistently present across all comparisons (Supplementary Material 1). Most proteins identified as significantly differential in BD were involved in the lipid metabolism, complement, and coagulation cascade (e.g., Apolipoproteins, Complement factors, and Coagulation factors).

**Table 2:**
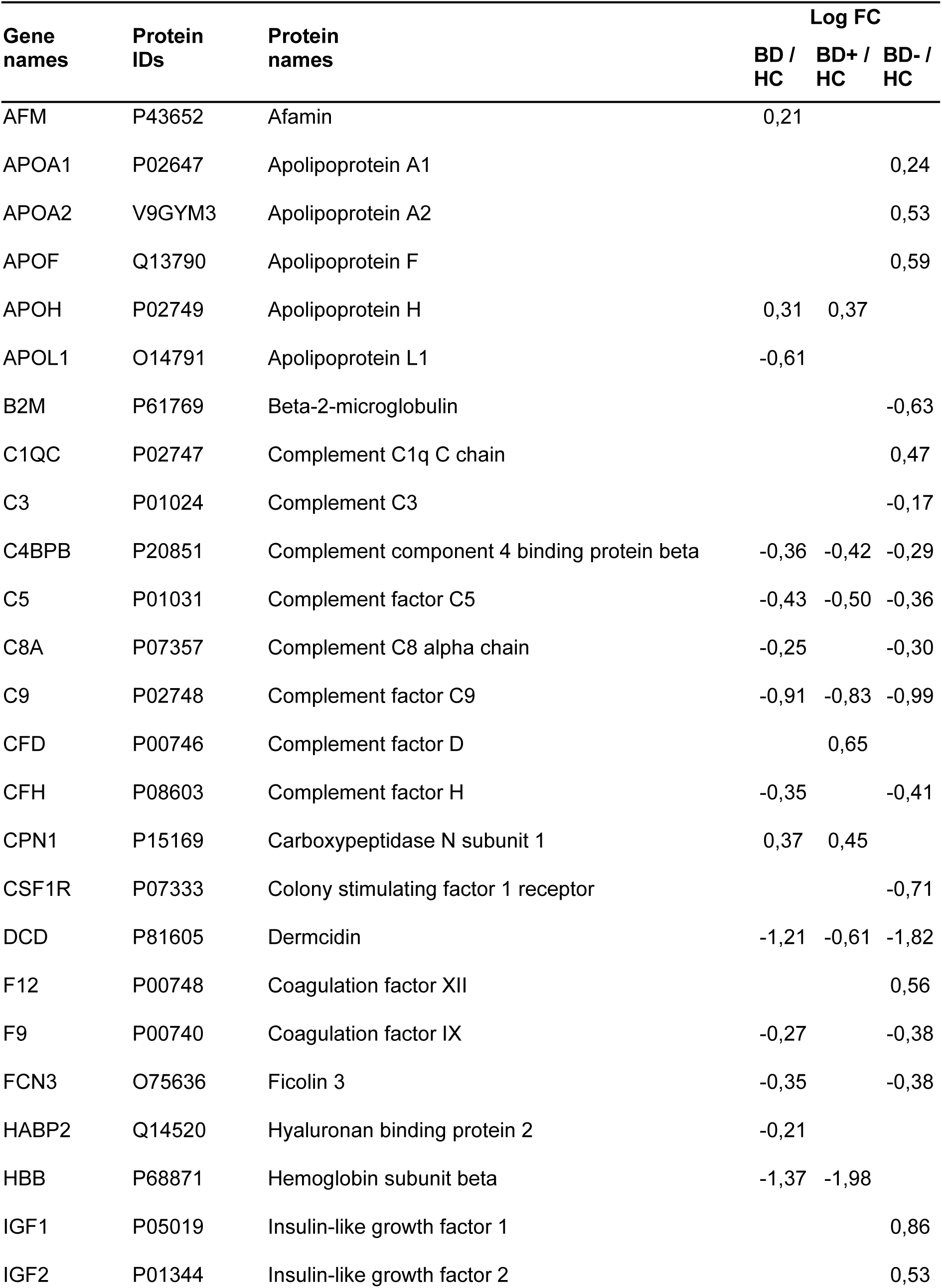
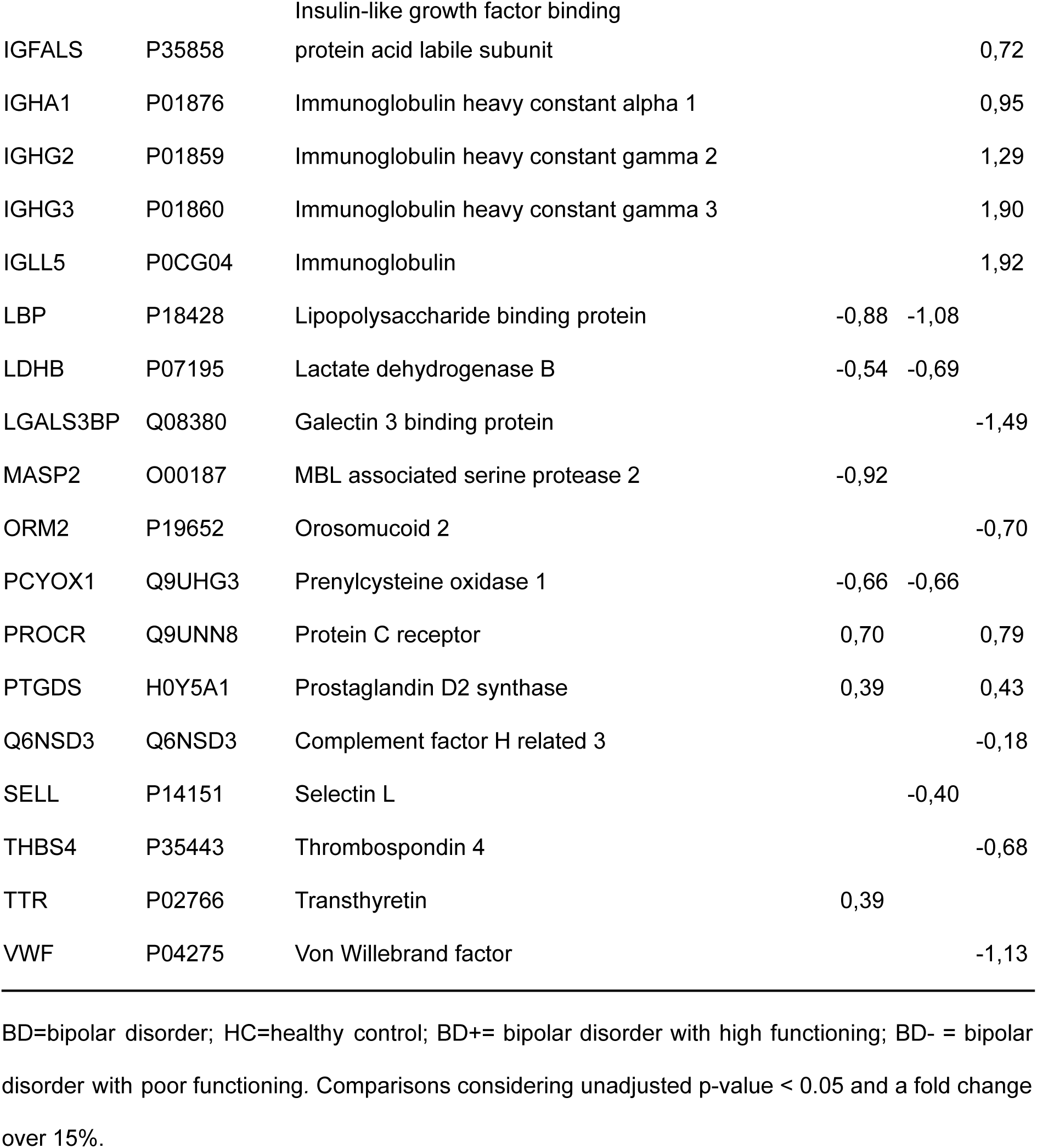
Molecular signatures from patients with BD and HC blood samples.

We utilized the Enrichr software [15] to detect KEGG pathways and diseases associated with BD. In all comparisons, the prominent signaling pathway identified through KEGG is the complement and coagulation cascade (Figure 2). Notably, the identified proteins in the BD-group were associated with cardiovascular diseases (Figure 3).

**Figure 2:**
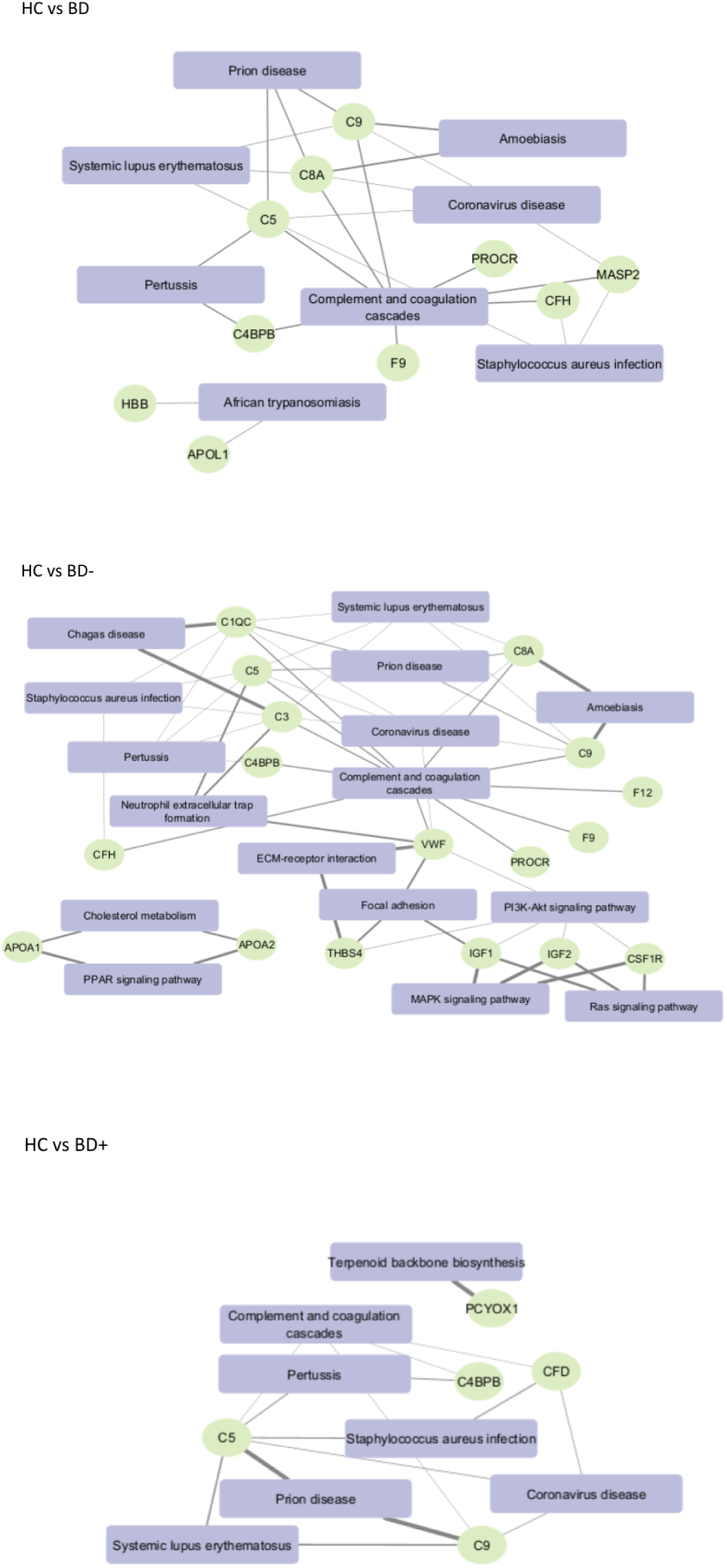
KEGG pathway analysis. Complement and coagulation cascade were the two main signaling pathways involved in BD, BD+ and BD-. Legend: HC = healthy control; BD = bipolar disorder; BD+ = bipolar disorder with high functioning; BD-= bipolar disorder with poor functioning. We considered an adjusted p-value < 0.00001 computed using the Benjamin-Hochberg method.

**Figure 3:**
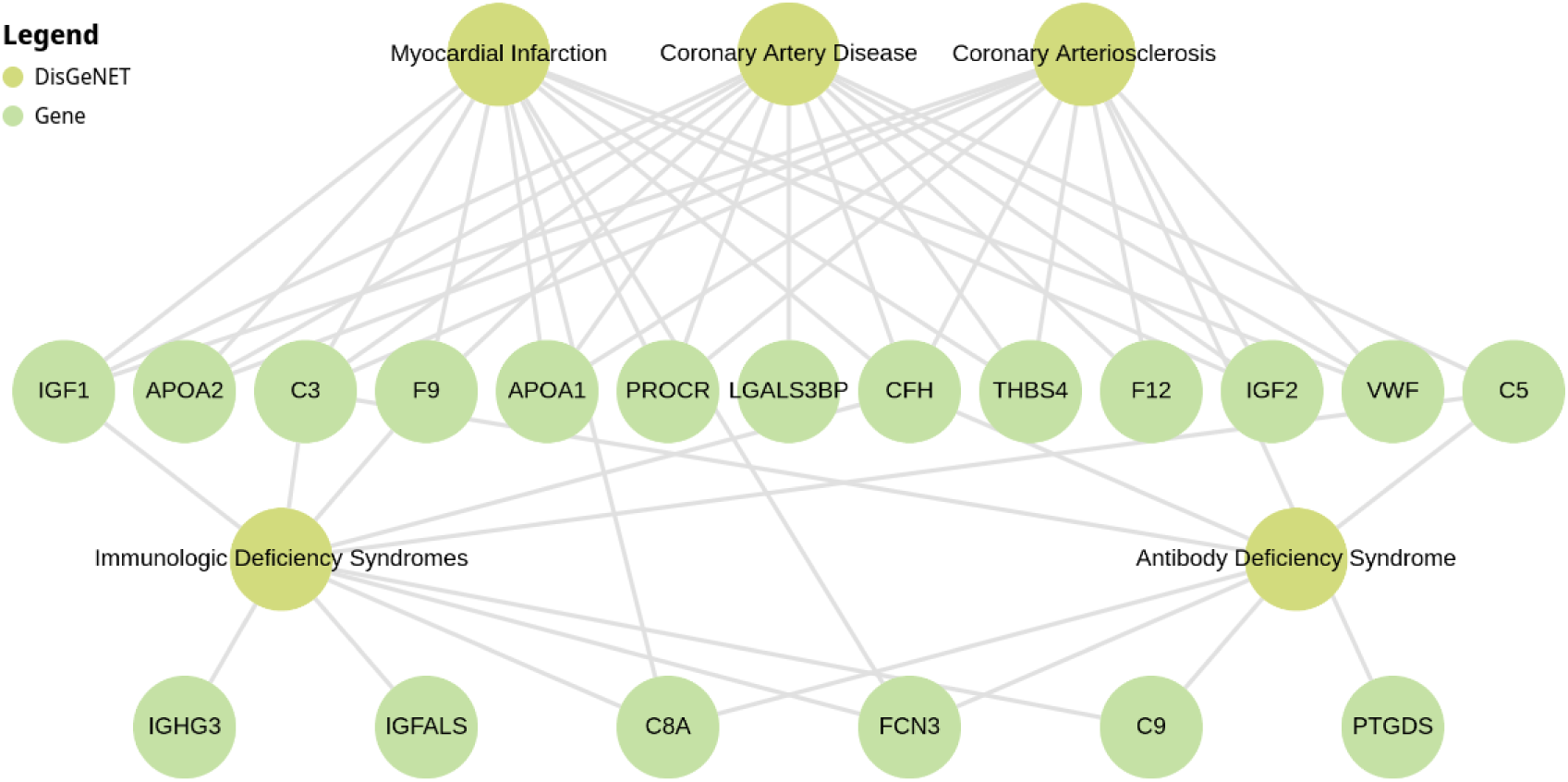
Enrichr analysis illustrating gene enrichment results using DisGeNET data. The figure highlights the association between BD-genes and cardiovascular diseases. Legend: BD-= bipolar disorder with poor functioning. We considered an adjusted p-value < 0.00001 computed using the Benjamin-Hochberg method.

WGCNA revealed a co-expression network of four separate modules within our sample of BD patients (Figure 4 A). We utilized the intramodular connectivity measure to designate the most strongly connected intramodular hub gene as the representative of the module. Demonstrating that intramodular hub genes exhibit a high correlation with the module eigengene. Among these 4 modules, we selected the “blue module” that presented a positive correlation with the BD group (r = 0.67) and a negative correlation with the HC group (r = - 0.67) (Figure 4 B and Figure 4 C). We did not observe a significant correlation between the blue module and studied clinical variables except for a positive correlation with the number of medications (r = 0.74) (Figure 4 D). A list of 36 proteins found within the blue module and relevant KEGG pathways was presented in Supplementary Material 2.

**Figure 4:**
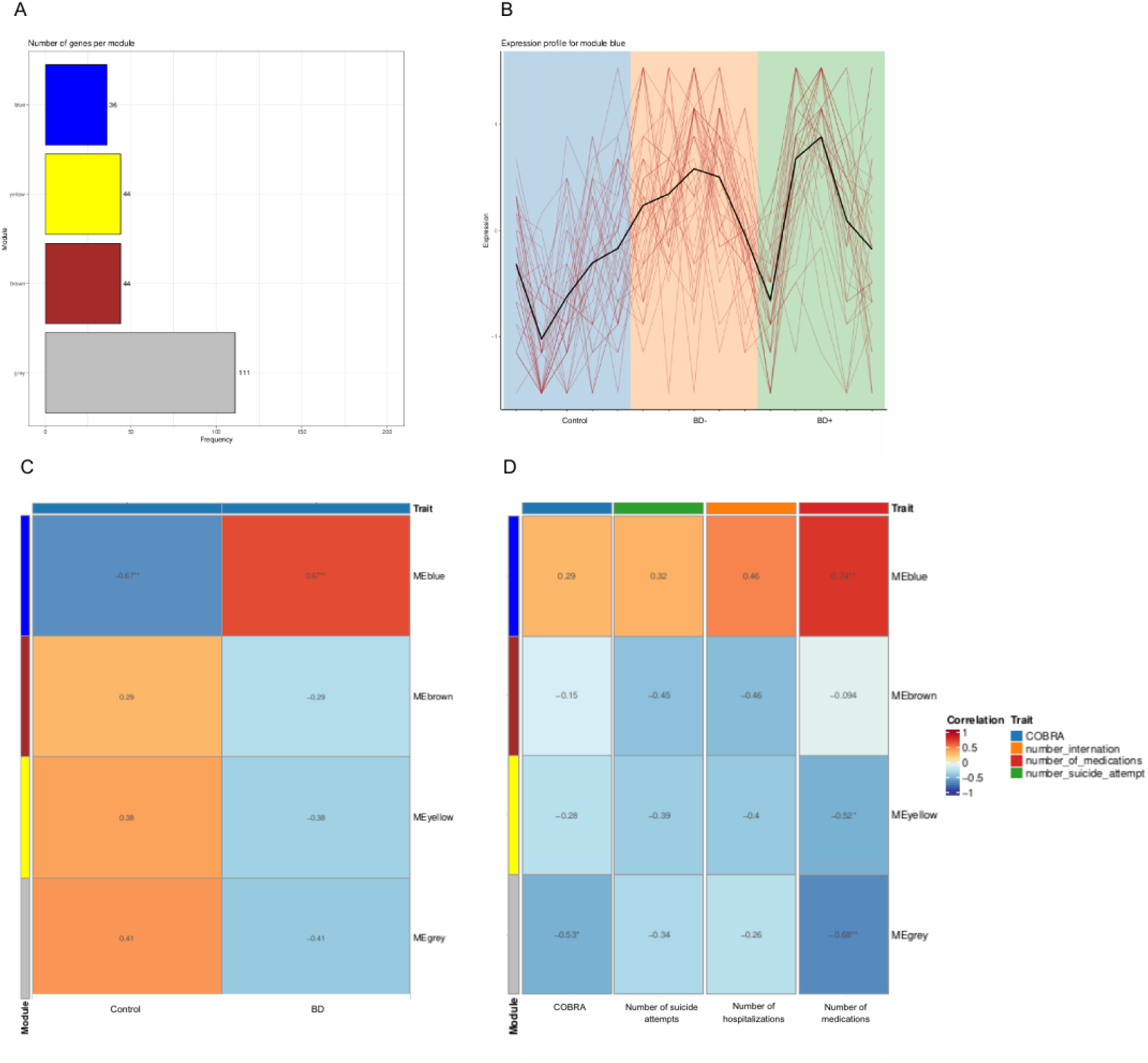
Weighted Gene Co-expression Network Analysis for the blue module. A) Number of proteins per module. B) A gene expression pattern in the HC group shows a negative distinction from the BD- and BD+ groups. C) Positive correlation between the blue module (y-axis) and the BD group (x-axis) and a negative correlation between the blue module and the HC group. The brown, yellow, and gray modules did not correlate significantly. D) The blue module exhibits a robust positive correlation with the quantity of medications our patients use. The remaining modules only display negative correlations with the covariates. Legend: BD = bipolar disorder; BD+ = bipolar disorder with high functioning; BD-= bipolar disorder with poor functioning; COBRA = Cognitive Bipolar Rating Assessment.

## 4. Discussion

Using a combined proteomic approach and bioinformatics, the current study revealed molecular signatures and pathways correlated with BD. Eight proteins exclusively characterized the molecular profile of patients with BD+ compared to HC, while 26 proteins, that is, three times more altered proteins, were observed in the BD-group. The altered proteins were mainly enriched in biological processes related to the complement system, coagulation cascade, lipid profile, and cardiovascular diseases; all these changes were most prominent in the BD-group. Thus, this is the first study to show, through mass spectrometry-based proteomic analysis, molecular patterns, pathways, and main medical diseases related to different stages of BD. Despite these encouraging discoveries, further research is warranted, encompassing larger sample cohorts and incorporating biological validation through the application of molecular biology methods.

Regarding biological processes and pathways, our findings support previous studies [16]–[19], indicating that the interplay between the complement system and coagulation cascade is involved in the etiology and progression of BD. The complement system originates from the serine protease reaction cascade, sharing a common ancestry with the coagulation factors encoded by the same ancestor genes [20], [21]. Beyond their common origin, these systems exhibit similar roles in promoting the initial defense against infections and tissue repair, potentially contributing to homeostasis or the onset of pathological conditions. Analogous to the complement system, the coagulation cascade is a tightly regulated and coordinated process culminating in clot formation, constituting the hemostasis system when combined with the fibrinolytic system and platelets. The coagulation cascade activation involves primary and secondary hemostasis, often coinciding with the activation of inflammatory mechanisms [22]. Primary hemostasis involves platelet activation and aggregation, leading to fibrin formation by thrombin. This event triggers an acute inflammatory response to control tissue damage, halt blood loss, and prevent microbial infection. During secondary hemostasis, plasmin dissolves fibrin and reparative inflammatory cells, collectively working to remodel and repair damaged tissue [23]. Under normal conditions, a tightly regulated hemostasis system poses minimal risk of complications or failed responses. However, dysregulation can lead to overactive platelet responses, abnormal fibrin formation, and impaired fibrinolysis, creating an environment conducive to cardiovascular events. Furthermore, the intercommunication between the complement and coagulation systems can amplify inflammatory processes within the vascular system, heightening the risk of cardiovascular events.

The differential proteins and biological processes identified in this study can represent systemic changes that occur with the progression of the disease or even the treatment response. In fact, the “blue module” identified by the WGCNA analysis showed a correlation between BD diagnosis and the number of medications. Polypharmacotherapy can be regarded as an indicator of disease complexity, suggesting the presence of various health conditions and a challenge in managing these conditions. We showed that medical comorbidities, in particular, cardiovascular diseases, were more preeminent in BD-suggesting this subgroup of patients may experience a more chronic course of the illness and poor prognosis. An interesting study showed that premature mortality in BD is linked to various causes, including myocardial infarction, diabetes, chronic obstructive pulmonary disease, pneumonia, and suicide [24]. Metabolic, respiratory, and cardiovascular diseases are among the top 10 causes of death worldwide and are notably highly prevalent comorbidities in BD. Therefore, medical commodities in individuals with BD can exacerbate the mortality disparity between them and the general population [25]. Many of these comorbidities are closely linked to the complement system [26], coagulation cascade [27], and lipid metabolism [28]. Thus, complement and coagulation systems and lipid metabolism may be potential mediators of BD’s early onset of medical comorbidities. These findings suggest that collaborative care interventions focusing on the treatment of these comorbidities can significantly decrease early death among individuals with BD [3].

Furthermore, we cannot rule out the possible effect of psychotropic medications on our results since all patients in this study were under pharmacological treatment. Mood stabilizers, as well as many atypical antipsychotics commonly used for the treatment of BD, can promote weight gain and, thus, the entire cascade of metabolic effects that lead to long-term cardiovascular complications. Nevertheless, it’s crucial to acknowledge that, despite the apparent heightened risk of cardiovascular diseases linked to numerous psychotropic medications, there is no conclusive evidence that these medications autonomously amplify the risk of cardiovascular disease in individuals with BD. In fact, some evidence opposes this hypothesis, suggesting that enhanced symptom control might facilitate improved health-related behaviors, increased utilization of health services, and, indirectly, better physical health [5].

A comprehensive understanding of the molecular correlates involving lipid metabolism, complement activation, and coagulation cascade abnormalities establishes an important link between cellular mechanisms and comorbidities clinical manifestations in BD. In summary, the proteins and signaling pathways identified contribute to an enhanced understanding of the biological mechanisms linking BD with its associated comorbidities, particularly those related to cardiovascular, respiratory, and metabolic diseases. Exploring these associations has the potential to guide targeted interventions, addressing not only the mental health dimensions of BD but also the fundamentals of comorbidities.

Some limitations should be considered when interpreting the results of this study. Firstly, the sample number was relatively small, and all patients were on pharmacological treatment. The polypharmacy introduces additional complexities that warrant thorough analysis since the pharmacological agents can directly influence the investigated biological responses and generate synergistic or antagonistic effects in signaling pathways and observed outcomes. Also, individual variability in treatment response is a significant factor, making it challenging to draw definitive conclusions about the associations between identified proteins and comorbid health conditions. Secondly, algorithms and analysis methods can significantly influence the results. Different algorithms may yield distinct interpretations from the same datasets, and the biological interpretation of results obtained through bioinformatic approaches can be challenging. Due to this, we partnered with an experienced bioinformatician to work with the data. Third, a proteomic profile was obtained from a blood sample, potentially not fully capturing the complex molecular changes associated with BD throughout the brain.

In conclusion, our exploratory analysis of peripheral blood in distinct stages of BD patients, through LC-MS/MS, identified proteins predominantly associated with lipid metabolism, the complement system, and the coagulation cascade. These findings may represent systemic alterations that occur with the progression of the illness and a possible link between BD and medical comorbidities. This comprehensive understanding provides valuable insights for targeted interventions, addressing mental health aspects and mechanisms driving comorbidities in BD.

## Data Availability

All data produced in the present study are available upon reasonable request to the authors.

## Acknowledgments

Researchers thank the Coordenação de Aperfeiçoamento de Pessoal de Nível Superior - Brasil (CAPES) - Finance Code 001, CNPq PQ 2019 productivity scholarship 302382/2019-4.

## Authors Approval

All authors have seen and approved the manuscript.

Supplementary Material 1: Protein intersection in comparison groups. Venn Diagram displaying proteins identified in BD, BD+, and BD-comparison groups.

Supplementary Material 2: Proteins identified in the blue module. A list of proteins found within the blue module and relevant KEGG pathways.

## References

[1] D. Vancampfort et al., “Physical activity and sedentary behavior in people with bipolar disorder: A systematic review and meta-analysis,” J Affect Disord, vol. 201, pp. 145–152, Sep. 2016, doi: 10.1016/j.jad.2016.05.020.

[2] C. Harris and B. Barraclough, “Excess mortality of mental disorder,” British Journal of Psychiatry, vol. 173, no. 1, pp. 11–53, Jul. 1998, doi: 10.1192/bjp.173.1.11.

[3] C. Crump, K. Sundquist, M. A. Winkleby, and J. Sundquist, “Comorbidities and Mortality in Bipolar Disorder,” JAMA Psychiatry, vol. 70, no. 9, p. 931, Sep. 2013, doi: 10.1001/jamapsychiatry.2013.1394.

[4] J. F. Hayes, J. Miles, K. Walters, M. King, and D. P. J. Osborn, “A systematic review and meta-analysis of premature mortality in bipolar affective disorder,” Acta Psychiatr Scand, vol. 131, no. 6, pp. 417–425, Jun. 2015, doi: 10.1111/acps.12408.

[5] B. I. Goldstein et al., “Call to action regarding the vascular-bipolar link: A report from the Vascular Task Force of the International Society for Bipolar Disorders,” Bipolar Disord, vol. 22, no. 5, pp. 440–460, Aug. 2020, doi: 10.1111/bdi.12921.

[6] T. B. Biazus et al., “All-cause and cause-specific mortality among people with bipolar disorder: a large-scale systematic review and meta-analysis,” Mol Psychiatry, vol. 28, no. 6, pp. 2508–2524, Jun. 2023, doi: 10.1038/s41380-023-02109-9.

[7] M. P. Vasconcelos-Moreno et al., “Telomere Length, Oxidative Stress, Inflammation and BDNF Levels in Siblings of Patients with Bipolar Disorder: Implications for Accelerated Cellular Aging,” International Journal of Neuropsychopharmacology, vol. 20, no. 6, pp. 445–454, Jun. 2017, doi: 10.1093/ijnp/pyx001.

[8] L. B. Rizzo et al., “The theory of bipolar disorder as an illness of accelerated aging: Implications for clinical care and research,” Neurosci Biobehav Rev, vol. 42, pp. 157–169, May 2014, doi: 10.1016/j.neubiorev.2014.02.004.

[9] B. Baykara et al., “Brain-derived neurotrophic factor in bipolar disorder: Associations with age at onset and illness duration,” Prog Neuropsychopharmacol Biol Psychiatry, vol. 108, p. 110075, Jun. 2021, doi: 10.1016/j.pnpbp.2020.110075.

[10] F. M. Lima et al., “Validity and reliability of the Cognitive Complaints in Bipolar Disorder Rating Assessment (COBRA) in Brazilian bipolar patients,” Trends Psychiatry Psychother, vol. 40, no. 2, pp. 170–178, Apr. 2018, doi: 10.1590/2237-6089-2017-0121.

[11] A. R. Rosa et al., “Clinical predictors of functional outcome of bipolar patients in remission,” Bipolar Disord, vol. 11, no. 4, pp. 401–409, Jun. 2009, doi: 10.1111/j.1399-5618.2009.00698.x.

[12] A. R. Rosa et al., “Clinical Staging in Bipolar Disorder,” J Clin Psychiatry, vol. 75, no. 05, pp. e450–e456, May 2014, doi: 10.4088/JCP.13m08625.

[13] A. R. Rosa et al., “Validity and reliability of the Functioning Assessment Short Test (FAST) in bipolar disorder,” Clinical Practice and Epidemiology in Mental Health, vol. 3, no. 1, p. 5, 2007, doi: 10.1186/1745-0179-3-5.

[14] P. Langfelder and S. Horvath, “WGCNA: an R package for weighted correlation network analysis,” BMC Bioinformatics, vol. 9, no. 1, p. 559, Dec. 2008, doi: 10.1186/1471-2105-9-559.

[15] E. Y. Chen et al., “Enrichr: interactive and collaborative HTML5 gene list enrichment analysis tool,” BMC Bioinformatics, vol. 14, no. 1, p. 128, Dec. 2013, doi: 10.1186/1471-2105-14-128.

[16] A. Reginia et al., “Assessment of Complement Cascade Components in Patients With Bipolar Disorder,” Front Psychiatry, vol. 9, Nov. 2018, doi: 10.3389/fpsyt.2018.00614.

[17] P. R. Ziani et al., “Potential Candidates for Biomarkers in Bipolar Disorder: A Proteomic Approach through Systems Biology,” Clinical Psychopharmacology and Neuroscience, vol. 20, no. 2, pp. 211–227, May 2022, doi: 10.9758/cpn.2022.20.2.211.

[18] E. C. Santa Cruz, F. da S. Zandonadi, W. Fontes, and A. Sussulini, “A pilot study indicating the dysregulation of the complement and coagulation cascades in treated schizophrenia and bipolar disorder patients,” Biochimica et Biophysica Acta (BBA) - Proteins and Proteomics, vol. 1869, no. 8, p. 140657, Aug. 2021, doi: 10.1016/j.bbapap.2021.140657.

[19] J. E. Rodrigues et al., “Systematic Review and Meta-Analysis on MS-Based Proteomics Applied to Human Peripheral Fluids to Assess Potential Biomarkers of Bipolar Disorder,” Int J Mol Sci, vol. 23, no. 10, p. 5460, May 2022, doi: 10.3390/ijms23105460.

[20] S. Oncul and V. Afshar-Kharghan, “The interaction between the complement system and hemostatic factors,” Curr Opin Hematol, vol. 27, no. 5, pp. 341–352, Sep. 2020, doi: 10.1097/MOH.0000000000000605.

[21] M. M. Krem and E. Di Cera, “Evolution of enzyme cascades from embryonic development to blood coagulation,” Trends Biochem Sci, vol. 27, no. 2, pp. 67–74, Feb. 2002, doi: 10.1016/S0968-0004(01)02007-2.

[22] J. P. Luyendyk, J. G. Schoenecker, and M. J. Flick, “The multifaceted role of fibrinogen in tissue injury and inflammation,” Blood, vol. 133, no. 6, pp. 511–520, Feb. 2019, doi: 10.1182/blood-2018-07-818211.

[23] R. Chaudhry, S. M. Usama, and H. M. Babiker, Physiology, Coagulation Pathways. 2023.

[24] B. Roshanaei-Moghaddam and W. Katon, “Premature Mortality From General Medical Illnesses Among Persons With Bipolar Disorder: A Review,” Psychiatric Services, vol. 60, no. 2, Feb. 2009, doi: 10.1176/appi.ps.60.2.147.

[25] D. Schoepf and R. Heun, “Comorbid medical illness in bipolar disorder,” British Journal of Psychiatry, vol. 206, no. 6, pp. 522–523, Jun. 2015, doi: 10.1192/bjp.206.6.522a.

[26] J. Østergaard, T. K. Hansen, S. Thiel, and A. Flyvbjerg, “Complement activation and diabetic vascular complications,” Clinica Chimica Acta, vol. 361, no. 1–2, pp. 10–19, Nov. 2005, doi: 10.1016/j.cccn.2005.04.028.

[27] H. ten Cate, T. M. Hackeng, and P. G. de Frutos, “Coagulation factor and protease pathways in thrombosis and cardiovascular disease,” Thromb Haemost, vol. 117, no. 07, pp. 1265–1271, Nov. 2017, doi: 10.1160/TH17-02-0079.

[28] C. W. Agudelo, G. Samaha, and I. Garcia-Arcos, “Alveolar lipids in pulmonary disease. A review,” Lipids Health Dis, vol. 19, no. 1, p. 122, Dec. 2020, doi: 10.1186/s12944-020-01278-8.

