## Supplementary figures and images for "Proteomic Insights into Biology of Bipolar Disorder: Implications for Health Complexity and Mortality"

### Supplementary Material 1

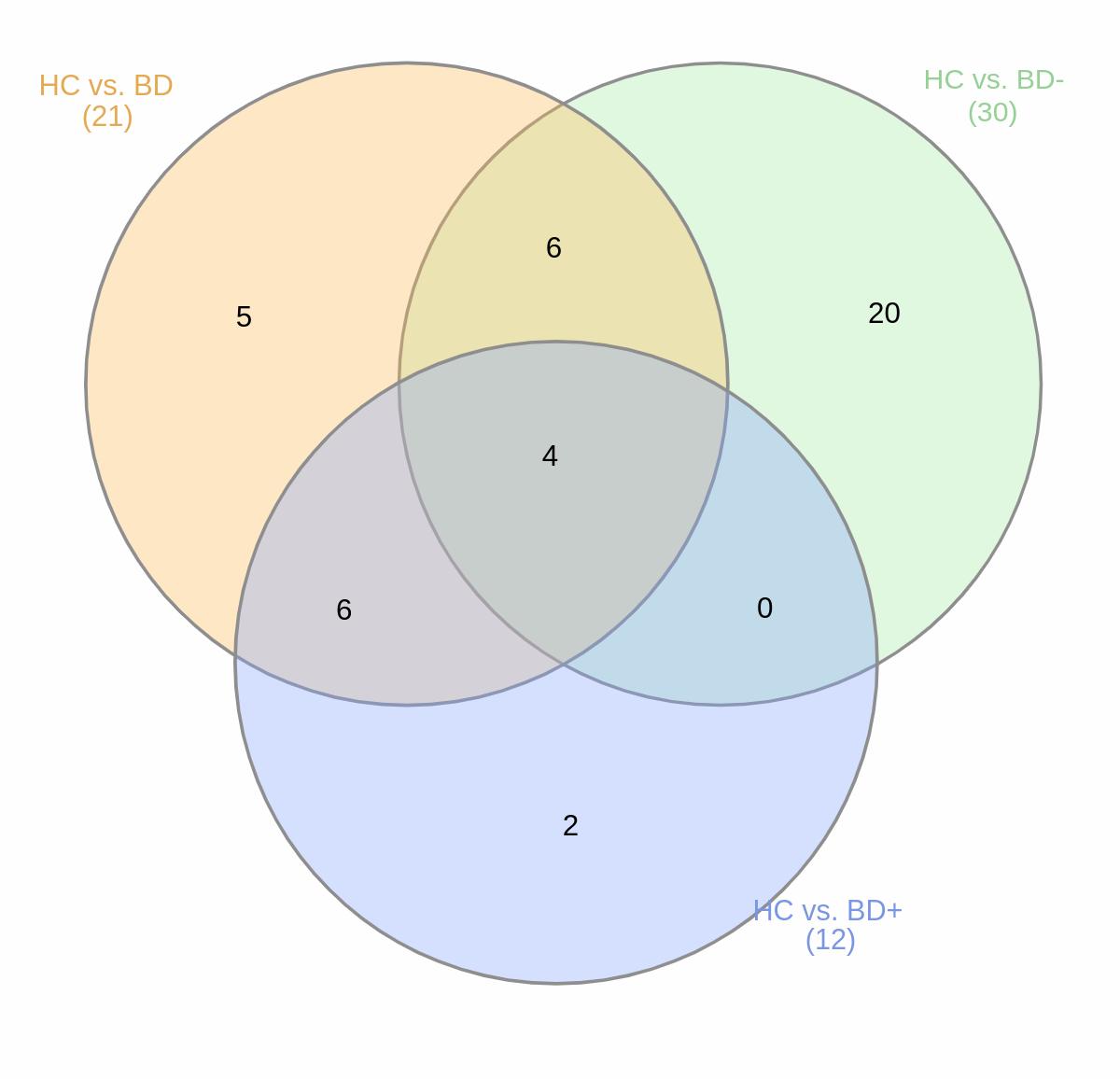
